# Proteomic associations with forced expiratory volume – a Mendelian randomisation study

**DOI:** 10.1101/2023.06.30.23292035

**Authors:** GT Axelsson, T Jonmundsson, YJ Woo, EA Frick, T Aspelund, JJ Loureiro, AP Orth, LL Jennings, G Gudmundsson, V Emilsson, V Gudmundsdottir, V Gudnason

## Abstract

A decline in forced expiratory volume (FEV1) is a hallmark of obstructive respiratory diseases, an important cause of morbidity among the elderly. While some data exist on biomarkers that are related to FEV1, we sought to do a systematic analysis of causal relations of biomarkers with FEV1.

Data from the general population-based AGES-Reykjavik study were used. Proteomic measurements were done using 4,782 DNA aptamers (SOMAmers). Data from 1,648 participants with spirometric data were used to assess the association of SOMAmer measurements with FEV1 using linear regression. Bi-directional Mendelian randomisation (MR) analyses were done to assess causal relations of observationally associated SOMAmers with FEV1, using genotype and SOMAmer data from 5,368 AGES-Reykjavik participants and genetic associations with FEV1 from a publicly available GWAS (n = 400,102).

In observational analyses, 473 SOMAmers were associated with FEV1 after multiple testing adjustment. The most significant were R-Spondin 4, Alkaline Phosphatase, Placental Like 2 and Retinoic Acid Receptor Responder 2. Of the 235 SOMAmers with genetic data, eight were associated with FEV1 in MR analyses. Three were directionally consistent with the observational estimate, Thrombospondin 2 (THBS2), Endoplasmic Reticulum Oxidoreductase 1 Beta and Apolipoprotein M. THBS2 was further supported by a colocalization analysis. Analyses in the reverse direction, testing whether changes in SOMAmer levels were caused by changes in FEV1, were performed but no significant associations were found after multiple testing adjustments.

In summary, this large scale proteogenomic analyses of FEV1 reveals protein markers of FEV1, as well as several proteins with potential causality to lung function.

## Introduction

Chronic respiratory diseases such as chronic obstructive pulmonary disease (COPD) are a leading global cause of mortality and morbidity, with their relative importance increasing in the last decades (1). Diagnosis of COPD is based on obstruction on pulmonary function testing, by a low forced expiratory volume in one second (FEV_1_) relative to the forced vital capacity (FVC), and a progressive decline in pulmonary function is a hallmark of the disease (2). While it is undisputed that exposure to external harmful stimuli such as cigarette smoke and biomass fumes are substantial risk factors for pulmonary function decline, intrinsic factors such as genetics and gene-environment interactions play a significant part as well (2–4). Genome wide association studies (GWAS) have found several genetic polymorphisms that are associated with COPD and lung function decline, including polymorphisms in or near genes encoding matrix metalloproteinase 12, nicotinic acetylcholine receptor, hedgehog interacting protein, glutathione S-transferase, C-terminal domain–containing protein (5–9) and the antiprotease alpha-1-antitrypsin (10). In addition to genetic polymorphisms, biomarkers that predict COPD or lung function have been discovered. Among them are inflammatory markers, soluble receptor for advanced glycoprotein end products (AGER), club cell secretory protein 16 (SCGB1A1) and surfactant protein D (SFTPD) (11–14). For several of these biomarkers, estimations of potential causality have been made. AGER, SCGB1A1 and SFTPD have been suggested to be causally associated with COPD or with lung function, while analyses have pointed against such an association for the inflammatory markers CRP and IL-6 (15–19). Still, while substantial epidemiologic data exist regarding single genetic markers and biomarkers that predict COPD or lung function, a systematic large-scale analysis of observational and causal associations of protein markers with lung function has not been undertaken to our knowledge.

Proteomics have emerged as a way of exploring biologic predictors of disease, especially with the advent of methods that allow for measurement and evaluation of thousands of proteins in biological samples from participants of large cohort studies (20). In addition to predicting disease and disease-related outcomes, integration of genetic data allows one to make assumptions regarding causality with proteomic markers. Mendelian randomization (MR) is such a method and utilizes genetic polymorphisms as instrumental variables to assess the relationship of an exposure with an outcome. As chromosomal alleles are randomly allocated during gamete formation, this methodology allows one to avoid the effect of confounders and to infer causality from epidemiologic data (21).

The aim of the study was to systematically explore the associations of a multitude of protein markers with lung function and then to assess the potential causal relationships of these protein markers with lung function by use of bi-directional Mendelian randomization.

## Methods

### Study phenotyping

The Age/Gene Environment Susceptibility (AGES)-Reykjavik study is a population-based cohort study of 5,764 elderly Icelanders that was carried out between 2002 and 2006. The participants, aged from 66 to 96 years (mean 76 years), were all prior participants of the Reykjavik Study done decades earlier. As part of AGES-Reykjavik, the participants underwent extensive phenotyping by questionnaires, physiological measurements, imaging studies and laboratory measurements, during a single clinic visit. The study was approved by the Icelandic National Bioethics Committee (VSN-00-063) and the Institutional Review Board of the Intramural Program of the National Institute for Aging, with informed consent obtained from all participants. Further details of the study design are previously published (22).

A subset of the study participants underwent lung function testing in a standardised manner. The device used was a Vitalograph Gold Standard Plus (Vitalograph Ltd., UK). Each participant completed three attempts. Participants with spirometry of acceptable quality were included (n = 1,660). Spirometry measures from participants that completed at least two attempts with a no more than 300 ml difference between the attempts and exhalation for at least 6 seconds were deemed acceptable (23). Smoking history was ascertained from questionnaires while anthropometric measurements were done during the clinic visit. Protein measurements were done in serum samples from participants using a high throughput proteomics technology, the SOMAscan (SomaLogic, Boulder, CO) in which DNA aptamers (Slow-Off Rate Modified Aptamers (SOMAmers)) bind to target protein epitopes and are then quantified with the help of fluorescence after wash-out of unbound aptamers and proteins. Measurements in AGES-Reykjavik were done with a 5,034 SOMAmer platform in serum from 5,457 AGES-Reykjavik participants, targeting 4,135 individual human proteins. For the analyses, 4,782 SOMAmers targeting human proteins were used. Measurement data were transformed using Box-Cox transformation and extreme outliers were excluded, as previously described (24).

### Statistical analyses

A flow chart of study design is shown in Figure 1. Descriptive statistics were compiled for participants with acceptable pulmonary function tests, demographic covariate data and protein measurements available (n = 1,648). Using data from these participants, the association of all measured human SOMAmers (n = 4,782) with FEV_1_ was assessed using linear regression modelling. These analyses were adjusted for variables that are commonly used to predict FEV_1_ in clinical practice, age, sex, height, age squared, and height squared (25). Adjustment for multiple testing was done using the Benjamini-Hochberg False Discovery Rate (FDR). To understand how tobacco smoking affected these associations, the analyses were repeated with participants stratified by smoking history (ever-smokers versus never-smokers). Genes encoding proteins significantly associated with the outcome after adjustment for multiple testing were subjected to over-representation analysis of Gene Ontology terms (26). In secondary analyses we explored the protein associations with additional outcomes, including FVC, FEV_1_/FVC ratio and COPD. Additional covariates included smoking status, pack years of smoking, weight and eGFR.

**Figure 1.**
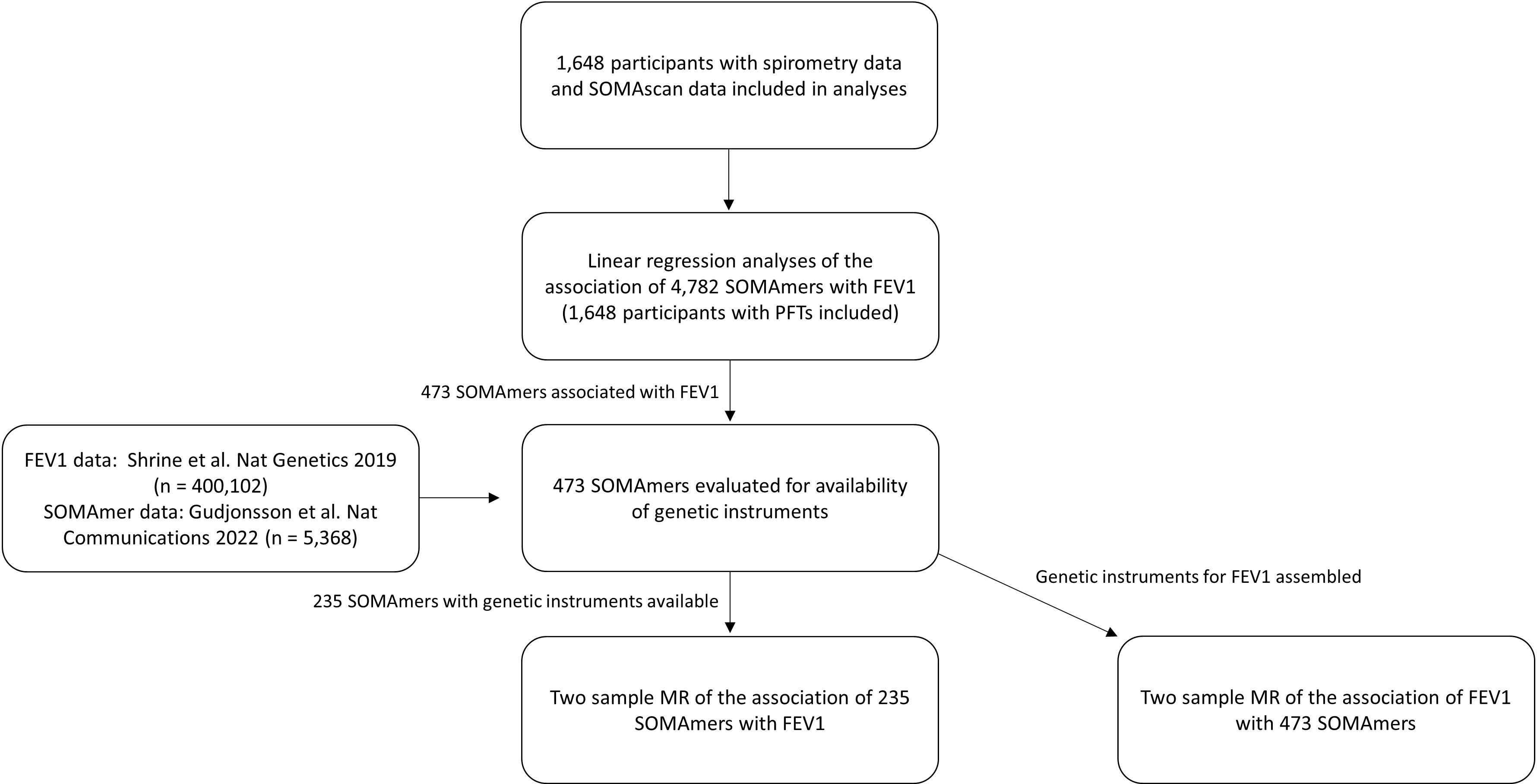
A flow chart of the study design

For SOMAmers that were associated with FEV_1_ after adjustment for multiple testing, a bi-directional two-sample MR analysis was performed for the association of SOMAmers with FEV_1_. MR analyses were done for SOMAmers that were observationally associated with FEV_1_ and had genetic instruments available. Genetic instruments for SOMAmers were defined as follows. The associations of single-nucleotide polymorphisms (SNPs) with SOMAmers were calculated using data from 5,368 participants of AGES-Reykjavik, as previously published and described in detail (24). The possible instrumental variables for each SOMAmer were defined as SNPs located within a cis-window near the gene coding for the protein measured by the SOMAmer, defined as within 500 kb up- and downstream of the gene. SNPs with a window-wide significant association (p < 0.05/number of SNPs in cis-window) with a given SOMAmer were considered as potential instruments. For each gene, SNPs were filtered based on linkage disequilibrium (LD; r^2^ < 0.2) or distance (>1 mb) using Plink v1.9. Instruments were considered valid if F > 10 for the association of instruments with SOMAmers. The associations of the instrumental variables with FEV_1_ were obtained from a GWAS of lung function in which data from two cohorts, UKBiobank and SpiroMeta Consortium, were meta-analysed. The total number of participants in that analysis was 400,102 (27). A MR analysis in the reverse direction was performed using all observationally associated SOMAmers as outcomes. Genetic instruments for FEV1 were defined based on the same GWAS on lung function as described above (27). For these analyses, SNPs associated with FEV_1_ (p < 5×10^−8^) were used as instruments after clumping within a cis-window 5 mb up- and downstream of the gene, by an LD threshold of 0.01. The associations of the instrumental variables with SOMAmer levels were obtained from AGES-Reykjavik data (24).

For the MR analyses, SNPs were harmonised using the TwoSampleMR R package. The MR estimates for FEV_1_ were obtained using the generalized weighted least squares (GWLS) method, accounting for correlation between instruments (28), except for instances where only one instrument was available, in which Wald ratios were calculated. For each MR analysis with more than two instruments, sensitivity analyses using the weighted median and Egger estimators were done to assess the validity of instruments and limit the effect of pleiotropic associations, respectively. Results were considered to pass these sensitivity analyses when the following conditions were met. For the weighted median estimator, the weighted median estimate had to be significant and directionally consistent with the GWLS estimate and for Egger, the Egger estimate had to be directionally consistent with the GWLS estimate and the intercept not significant. In analyses in the reverse direction, estimates were obtained using the inverse variance weighted method. Adjustment for multiple testing was done using the Benjamini-Hochberg False Discovery Rate (FDR).

A colocalization analysis was performed to provide additional causal support for analytes associated with FEV1 in MR analyses. In that aim, AGES-Reykjavik serum pQTLs (24), summary statistics for FEV1 (29) and plasma pQTLs from deCODE (30) were harmonized to account for strand orientation and differences in genome builds. All studies were lifted over to build GRCh38 for colocalization when needed. For the QTLs, only putative cis-regulatory variants were examined defined by 500kb away from the gene body. All regions with variants that had associations of P < 1×10^−5^ were fine-mapped using Sum of Single Effects (SuSiE) (31) based on 1000 genome population reference (32). 95% credible-sets were filtered and colocalization between GWAS and QTLs was performed using fastENLOC using the posterior inclusion probabilities estimated in SuSiE (31, 33). Regional colocalization probability (RCP) was calculated by summing the colocalization probability within each 95% credible-set and filtered colocalization results at RCP > 80% for further examination. RCP represents the probability that a given genomic region contains a single colocalized variant (34). Visualization was done using Julia 1.7 and libraries within (35).

## Results

### The protein profile of pulmonary function

Descriptive statistics for participants that had pulmonary function testing data are shown in Table 1. A majority of participants were female (56%) and participants were on average 76 years old. Most participants had a history of smoking (59%).

**Table 1.** Overview of study participants

| n | 1648 |
| --- | --- |
| Sex = Female (%) | 917 (55.6) |
| Age (mean (SD)) | 76.28 (5.70) |
| Height (mean (SD)) | 167.22 (9.48) |
| Weight (mean (SD)) | 75.61 (14.67) |
| BMI (mean (SD)) | 26.97 (4.38) |
| Eversmoker (%)<sup>†</sup> | 969 (59.4) |
| FEV<sub>1</sub> (mean (SD)) | 2.12 (0.69) |
| FEV<sub>1</sub> % of predicted (mean (SD))* | 0.89 (0.21) |
| FVC (mean (SD)) | 2.97 (0.83) |
| FVC % of predicted (mean (SD))* | 0.92 (0.17) |
| FEV<sub>1</sub>/FVC < 70% (%) | 578 (35.1) |

<sup>†</sup> 17 participants had missing smoking data.
\* Predicted values based on Hankinson et al. (25)

Of the 4,782 SOMAmers tested, 473 were observationally associated with FEV_1_ after adjustment for multiple testing (Table 2, Table S1, Figure 2). The most significantly associated SOMAmers measured were R-Spondin 4 (RSPO4, β = −0.099, p = 1.13×10^−13^), Alkaline Phosphatase, Placental Like 2 (ALPPL2, β = −0.084, p = 4.76×10^−11^), Retinoic Acid Receptor Responder 2 (RARRES2, β = −0.093, p = 7.03×10^−11^), Sushi, Von Willebrand Factor Type A, EGF And Pentraxin Domain Containing 1 (SVEP1, β = −0.085, p = 1.67×10^−10^) and Polymeric Immunoglobulin Receptor (PIGR, β = −0.080, 1.80×10^−10^). Results for proteins that have been previously suggested as biomarkers of FEV_1_ (11) are shown in Table S2. Prior associations of SFTPD (beta = −0.05, p = 0.0002), CRP (beta = −0.06, p = 1.2×10^−7^), fibrinogen (beta = −0.04, p = 0.0008 for the stronger associated SOMAmer), IL6 (beta = −0.05, p = 9.6×10^−5^ for the stronger associated SOMAmer) and eotaxin (beta = −0.05, p = 0.0002) were reproduced in the AGES-Reykjavik data while associations for other suggested biomarkers of FEV_1_ were not.

**Figure 2.**
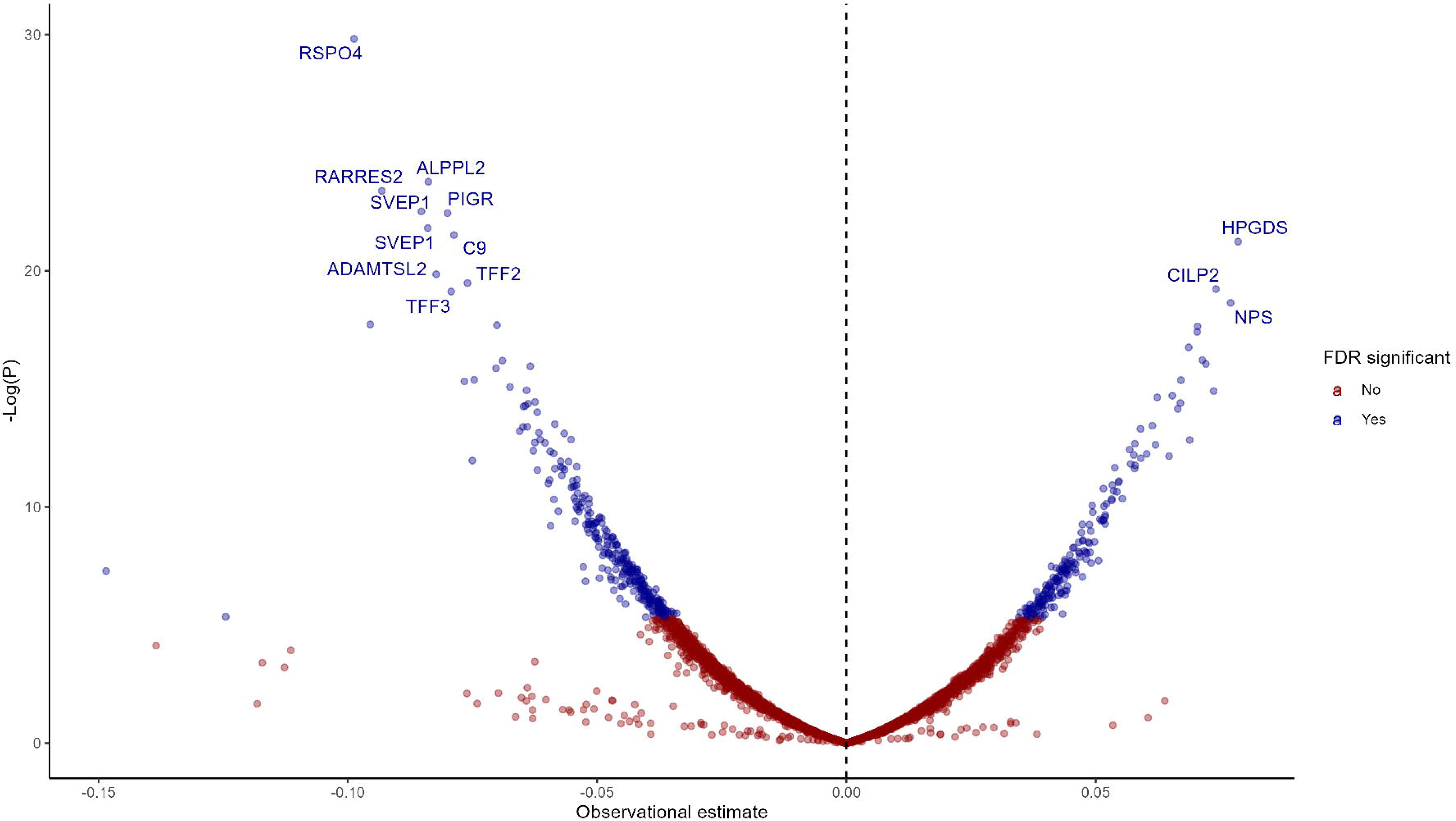
A volcano plot showing the observational associations of all 4782 SOMAmers with FEV1

**Table 2.** Observational associations of highly-associated proteins with FEV_1_

| SOMA | EGS | $\beta$ | 95% CI | P | FDR P |
| --- | --- | --- | --- | --- | --- |
| 8464_31_3 | RSPO4 | -0.099 | -0.125 - -0.073 | $1.13 \times 10^{-13}$ | $5.39 \times 10^{-10}$ |
| 7813_6_3 | ALPPL2 | -0.084 | -0.109 - -0.059 | $4.76 \times 10^{-11}$ | $1.12 \times 10^{-7}$ |
| 3079_62_2 | RARRES2 | -0.093 | -0.121 - -0.065 | $7.03 \times 10^{-11}$ | $1.12 \times 10^{-7}$ |
| 11178_21_3 | SVEP1 | -0.085 | -0.111 - -0.059 | $1.67 \times 10^{-10}$ | $1.72 \times 10^{-7}$ |
| 3216_2_2 | PIGR | -0.08 | -0.104 - -0.056 | $1.80 \times 10^{-10}$ | $1.72 \times 10^{-7}$ |
| 11109_56_3 | SVEP1 | -0.084 | -0.11 - -0.058 | $3.39 \times 10^{-10}$ | $2.70 \times 10^{-7}$ |
| 2292_17_4 | C9 | -0.079 | -0.103 - -0.054 | $4.55 \times 10^{-10}$ | $3.11 \times 10^{-7}$ |
| 12549_33_3 | HPGDS | 0.079 | 0.054 - 0.103 | $6.02 \times 10^{-10}$ | $3.60 \times 10^{-7}$ |
| 6379_62_3 | ADAMTSL2 | -0.082 | -0.109 - -0.055 | $2.40 \times 10^{-9}$ | $1.27 \times 10^{-6}$ |
| 9191_8_3 | TFF2 | -0.076 | -0.101 - -0.051 | $3.47 \times 10^{-9}$ | $1.66 \times 10^{-6}$ |
| 8841_65_3 | CILP2 | 0.074 | 0.049 - 0.099 | $4.47 \times 10^{-9}$ | $1.95 \times 10^{-6}$ |
| 8323_163_3 | TFF3 | -0.079 | -0.106 - -0.053 | $4.98 \times 10^{-9}$ | $1.99 \times 10^{-6}$ |
| 6390_18_3 | NPS | 0.077 | 0.051 - 0.103 | $8.04 \times 10^{-9}$ | $2.96 \times 10^{-6}$ |
| 2211_9_6 | TIMP1 | -0.096 | -0.129 - -0.062 | $2.00 \times 10^{-8}$ | $6.50 \times 10^{-6}$ |
| 13722_105_3 | C9 | -0.07 | -0.094 - -0.046 | $2.06 \times 10^{-8}$ | $6.50 \times 10^{-6}$ |
| 6605_17_3 | IGFALS | 0.07 | 0.046 - 0.095 | $2.18 \times 10^{-8}$ | $6.50 \times 10^{-6}$ |
| 12707_26_3 | DPYSL3 | 0.07 | 0.046 - 0.095 | $2.75 \times 10^{-8}$ | $7.72 \times 10^{-6}$ |
| 5728_60_3 | FCRL1 | 0.069 | 0.044 - 0.093 | $5.29 \times 10^{-8}$ | $1.40 \times 10^{-5}$ |
| 6075_61_3 | HEXB | 0.071 | 0.045 - 0.097 | $9.08 \times 10^{-8}$ | $2.22 \times 10^{-5}$ |
| 4496_60_2 | MMP12 | -0.069 | -0.094 - -0.044 | $9.28 \times 10^{-8}$ | $2.22 \times 10^{-5}$ |
| 6225_3_3 | PRSS8 | 0.072 | 0.046 - 0.099 | $1.07 \times 10^{-7}$ | $2.43 \times 10^{-5}$ |
| 4337_49_2 | CRP | -0.063 | -0.087 - -0.04 | $1.18 \times 10^{-7}$ | $2.57 \times 10^{-5}$ |
| 10620_21_3 | MSMB | -0.07 | -0.096 - -0.044 | $1.28 \times 10^{-7}$ | $2.67 \times 10^{-5}$ |
| 2609_59_2 | CST3 | -0.075 | -0.103 - -0.047 | $2.09 \times 10^{-7}$ | $4.05 \times 10^{-5}$ |
| 8885_6_3 | CACNA2D3 | 0.067 | 0.042 - 0.092 | $2.11 \times 10^{-7}$ | $4.05 \times 10^{-5}$ |

Results of linear regression analyses adjusted for sex, age, age squared, height and height squared.
Results are shown for the proteins with the most significant associations.

The proteins most significantly associated with FEV1 overall also had strong associations with FEV1 among ever-smokers, such as RSPO4 (β = −0.106, p = 4.83×10^−9^), RARRES2 (β = −0.11, p = 1.19×10^−8^) and ALPPL2 (β = −0.092, p = 5.73×10^−8^). Trefoil Factor 2 (TFF2, β = −0.095, p = 3.84×10^−8^), Complement 9 (C9, β = −0.095, p = 5.65×10^−8^) and TIMP Metallopeptidase Inhibitor 1 (TIMP1, β = −0.122, p = 1.18×10^−7^) also had notably strong associations with the outcome. However, associations of these proteins were much weaker among never-smokers, with the strongest associations observed for two SOMAmers measuring SVEP1 (β = −0.081, p = 4.52×10^−6^ for the stronger associated SOMAmer) (Table S3) in this subgroup. The proteins associated with FEV1 were most strongly enriched for Gene Ontology (GO) terms related to humoral immune response, regulation of neurogenesis, compliment activation and extracellular matrix organization (Figure S1, Table S4).

Secondary analyses showed a large overlap between the protein profile of FEV_1_ compared to that of FVC, FEV_1_/FVC ratio and COPD, with some additional proteins detected (204 significant SOMAmers in primary analysis were among 497 significant SOMAmers in secondary analyses; Tables S8-S15).

### Mendelian randomization analysis

Of 473 SOMAmers associated with FEV_1_, 235 (50%) had genetic instruments available and were included in MR analyses of the association of SOMAmers with FEV_1_. The instruments are shown in Table S5 for significant SOMAmers. Of the 235 SOMAmers tested, eight were significantly associated with FEV_1_ in MR analyses (Table 3), suggesting they may have a causal effect on lung function. Of those, three, Thrombospondin 2 (THBS2, β = −0.037, p = 9.53×10^−5^), Endoplasmic Reticulum Oxidoreductase 1 Beta (ERO1B, β = −0.025, p = 8.05×10^−4^) and Apolipoprotein M (APOM, β = 0.053, p = 9.72×10^−4^) were directionally consistent with the observational analyses. The other five SOMAmers with significant causal estimates, R-Spondin-2 (RSPO2), TIMP Metallopeptidase Inhibitor 4 (TIMP4), CD14, Heparin Binding Growth Factor (HDGF) and Parkinsonism Associated Deglycase (PARK7) were directionally inconsistent. Table 3 and Figure 4 show the 29 SOMAmers that were nominally associated with FEV_1_ (unadjusted p < 0.05) using MR and passed weighted median sensitivity testing. Among nominally associated SOMAmers, the directional consistency between causal and observational estimates was low, or 38%, suggesting that the potentially causal effects of the protein are generally not reflected in the observational estimates. Of the five previously suggested biomarkers of FEV_1_ listed in Table S2 that were observationally associated with FEV_1_, only three proteins had genetic instruments available, the acute phase reactants CRP and fibrinogen as well as eotaxin. None of these proteins were significantly associated with FEV_1_ in MR analyses (Table S6).

**Figure 3.**
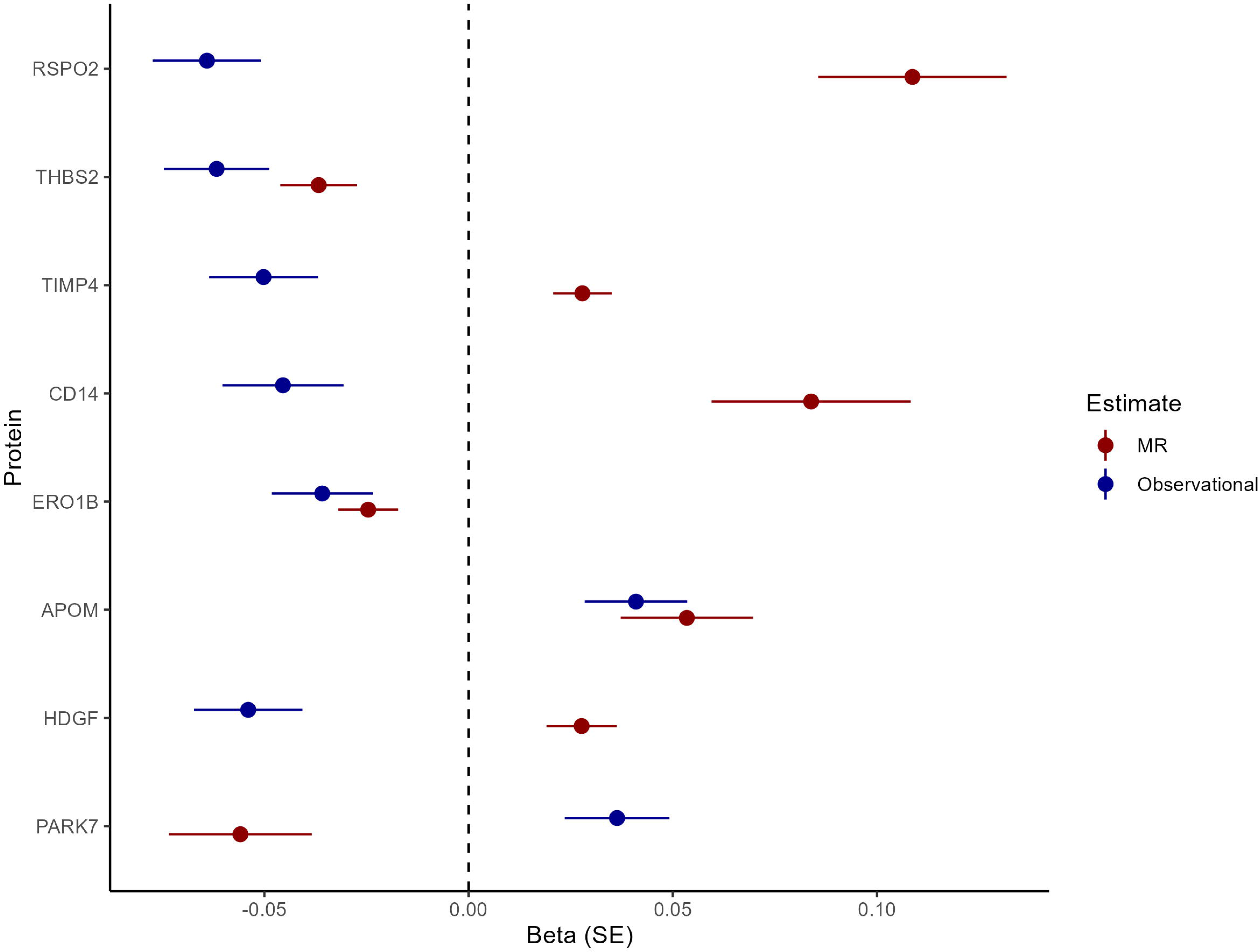
A forest plot showing the observational and mendelian randomisation estimates for FEV1 for causally associated SOMAmers

**Figure 4.**
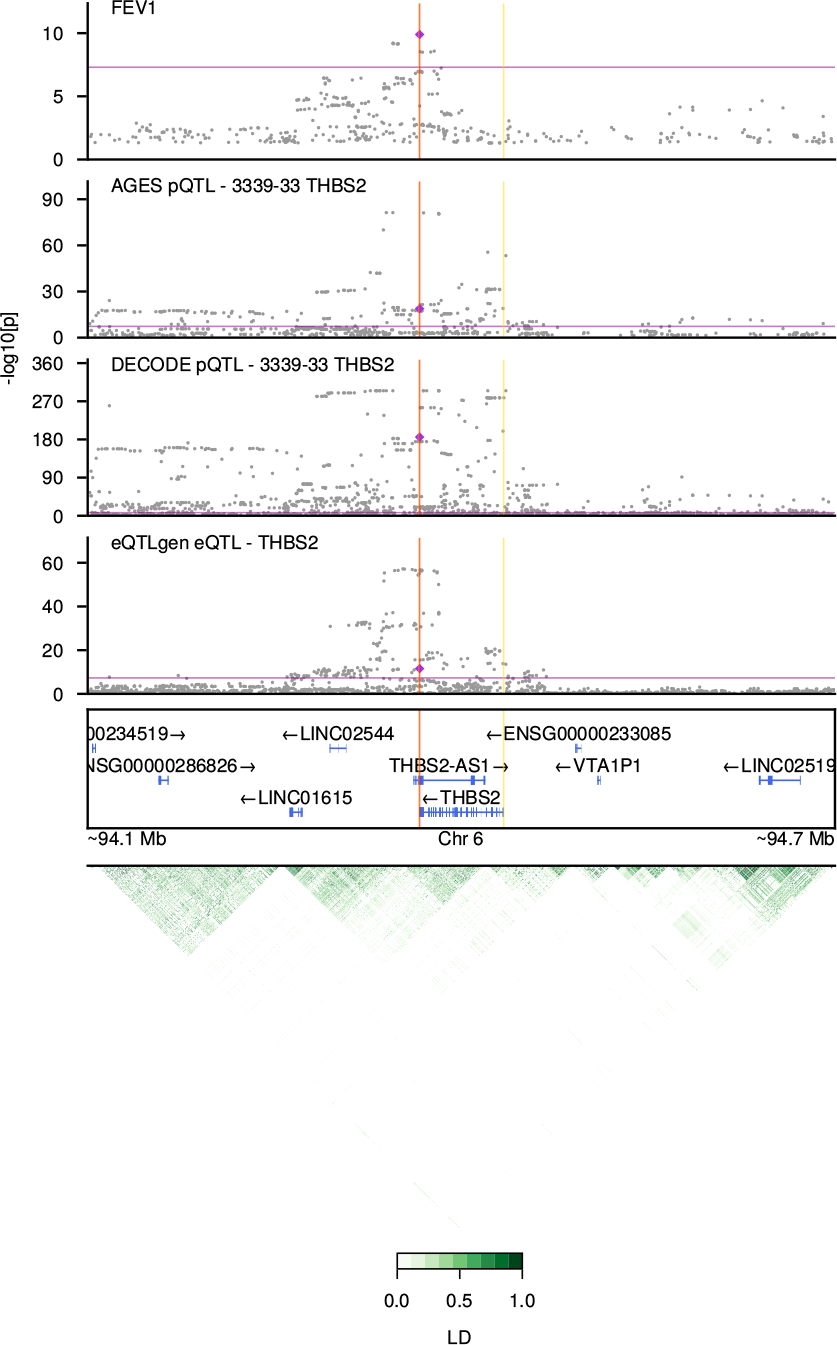
Colocalization plot for *THBS2*. FEV1 GWAS (29), AGES pQTL (24), and DECODE pQTL (30) data are shown from top to bottom. Grey circles are individual SNPs from each study. Both purple diamond and red vertical line represent the lead variant (rs3253, 3’ UTR variant). X-axis is genomic position within chromosome 6 and y-axis is-log10 transformed P-values. Purple horizontal lie delineates GWAS threshold at 5×10^−8^ and yellow vertical lines represent gene boundary for *THBS2*. Visualization is restricted to 150,000 bp upstream and downstream of THBS2. Linkage disequilibrium (r^2^) within the region is plotted at the bottom in green.

**Table 3.** Results of Mendelian randomisation analysis of the association of proteins with FEV_1_

| SOMA | Protein | nSNP | B - MR | SE | P - MR | FDR P - MR | Egger | B - Obs | P - Obs | FDR P - Obs |
| --- | --- | --- | --- | --- | --- | --- | --- | --- | --- | --- |
| **8409_3_3** | **RSPO2** | **3** | **0.109** | **0.023** | **2.44×10<sup>-6</sup>** | **5.73×10<sup>-4</sup>** | Yes | **-0.064** | **1.52×10<sup>-6</sup>** | **1.75×10<sup>-4</sup>** |
| **3339_33_1** | **THBS2** | **24** | **-0.037** | **0.009** | **9.53×10<sup>-5</sup>** | **7.85×10<sup>-3</sup>** | Yes | **-0.062** | **1.96×10<sup>-6</sup>** | **2.09×10<sup>-4</sup>** |
| **6462_12_3** | **TIMP4** | **17** | **0.028** | **0.007** | **1.00×10<sup>-4</sup>** | **7.85×10<sup>-3</sup>** | Yes | **-0.05** | **1.70×10<sup>-4</sup>** | **5.29×10<sup>-3</sup>** |
| **8969_49_3** | **CD14** | **6** | **0.084** | **0.024** | **5.85×10<sup>-4</sup>** | **0.034** | Yes | **-0.045** | **2.21×10<sup>-3</sup>** | **0.029** |
| **7994_41_3** | **ERO1B** | **12** | **-0.025** | **0.007** | **8.05×10<sup>-4</sup>** | **0.038** | Yes | **-0.036** | **3.78×10<sup>-3</sup>** | **0.042** |
| **14125_5_3** | **APOM** | **1** | **0.053** | **0.016** | **9.72×10<sup>-4</sup>** | **0.038** | - | **0.041** | **1.12×10<sup>-3</sup>** | **0.018** |
| **8953_47_3** | **HDGF** | **7** | **0.028** | **0.009** | **1.26×10<sup>-3</sup>** | **0.041** | Yes | **-0.054** | **5.10×10<sup>-5</sup>** | **2.26×10<sup>-3</sup>** |
| **9845_33_3** | **PARK7** | **2** | **-0.056** | **0.017** | **1.40×10<sup>-3</sup>** | **0.041** | - | **0.036** | **4.63×10<sup>-3</sup>** | **0.048** |
| 2668_70_2 | CAPN1 | 3 | -0.051 | 0.016 | 1.94×10<sup>-3</sup> | 0.051 | Yes | 0.04 | 2.93×10<sup>-3</sup> | 0.035 |
| 3290_50_2 | CD109 | 49 | -0.013 | 0.005 | 4.91×10<sup>-3</sup> | 0.111 | Yes | 0.044 | 4.23×10<sup>-4</sup> | 0.01 |
| 2797_56_2 | APOB | 9 | -0.029 | 0.011 | 5.18×10<sup>-3</sup> | 0.111 | Yes | 0.054 | 2.39×10<sup>-5</sup> | 1.28×10<sup>-3</sup> |
| 3580_25_8 | SERPINA1 | 9 | -0.027 | 0.01 | 6.03×10<sup>-3</sup> | 0.118 | Yes | -0.055 | 3.11×10<sup>-5</sup> | 1.61×10<sup>-3</sup> |
| 12395_86_3 | DARS2 | 2 | -0.058 | 0.022 | 9.64×10<sup>-3</sup> | 0.164 | - | -0.037 | 3.09×10<sup>-3</sup> | 0.036 |
| 5939_42_3 | TNFSF12 | 77 | 0.011 | 0.004 | 0.01 | 0.164 | Yes | 0.037 | 3.33×10<sup>-3</sup> | 0.038 |
| 8427_118_3 | RSPO3 | 17 | 0.033 | 0.013 | 0.01 | 0.164 | Yes | -0.06 | 1.68×10<sup>-5</sup> | 9.92×10<sup>-4</sup> |
| 2630_12_2 | IL1RAP | 58 | 0.006 | 0.002 | 0.018 | 0.259 | No | 0.037 | 3.38×10<sup>-3</sup> | 0.038 |
| 5070_76_3 | TNFRSF6B | 3 | 0.174 | 0.075 | 0.02 | 0.283 | Yes | -0.044 | 3.14×10<sup>-4</sup> | 8.25×10<sup>-3</sup> |
| 8992_1_3 | TMEM2 | 1 | -0.052 | 0.023 | 0.025 | 0.312 | - | 0.043 | 5.41×10<sup>-4</sup> | 0.012 |
| 8288_27_3 | APOH | 5 | -0.032 | 0.014 | 0.025 | 0.312 | Yes | 0.044 | 7.37×10<sup>-4</sup> | 0.014 |
| 13130_150_3 | HK2 | 2 | -0.04 | 0.019 | 0.032 | 0.338 | - | -0.042 | 8.84×10<sup>-4</sup> | 0.015 |
| 9870_17_3 | WARS | 5 | -0.021 | 0.01 | 0.032 | 0.338 | Yes | 0.041 | 1.31×10<sup>-3</sup> | 0.02 |
| 4297_62_3 | SPON1 | 8 | -0.026 | 0.012 | 0.032 | 0.338 | Yes | -0.052 | 1.17×10<sup>-4</sup> | 3.93×10<sup>-3</sup> |
| 2789_26_2 | MMP7 | 8 | 0.02 | 0.009 | 0.033 | 0.338 | Yes | -0.054 | 2.53×10<sup>-5</sup> | 1.34×10<sup>-3</sup> |
| 9172_69_3 | MMP8 | 13 | -0.014 | 0.007 | 0.036 | 0.343 | Yes | -0.041 | 1.03×10<sup>-3</sup> | 0.017 |
| 5646_20_3 | RNASE6 | 33 | 0.008 | 0.004 | 0.037 | 0.343 | Yes | -0.048 | 1.95×10<sup>-4</sup> | 5.80×10<sup>-3</sup> |
| 4546_27_3 | ADGRE2 | 29 | 0.012 | 0.006 | 0.039 | 0.343 | Yes | 0.057 | 7.36×10<sup>-6</sup> | 5.41×10<sup>-4</sup> |
| 3449_58_2 | SERPINA4 | 56 | -0.007 | 0.004 | 0.039 | 0.343 | Yes | 0.04 | 1.42×10<sup>-3</sup> | 0.021 |
| 10746_24_3 | DKK3 | 6 | -0.031 | 0.015 | 0.044 | 0.356 | Yes | 0.049 | 2.10×10<sup>-4</sup> | 6.09×10<sup>-3</sup> |
| 12387_7_3 | PDLIM4 | 11 | -0.025 | 0.012 | 0.044 | 0.356 | Yes | 0.046 | 5.08×10<sup>-4</sup> | 0.011 |

Shown are data for SOMAmers that had nominally significant associations using Mendelian randomisation and passed weighted median sensitivity analyses. In the column with results from Egger analyses, “Yes” denotes passing the sensitivity test, “No” denotes failing and “-” denotes insufficient number of SNPs to perform the test. Observational data are adjusted for sex, age, age squared, height and height squared.
**Bold** = Significantly associated with FEV<sub>1</sub> in MR analyses after multiple testing correction.
*Italic* = Consistent in direction between MR analyses and observational analyses

The 29 proteins identified from the MR analysis (Table 3) were further examined for colocalization evidence. Among the 8 proteins with significant MR association, a 3’ UTR variant (rs3253) from the FEV1 GWAS colocalized with THBS2 protein expression in AGES-Reykjavik (P = 1.4×10^−19^, β = −0.18, RCP=0.9976). The same variant was associated with THBS2 protein expression in the deCODE cohort (P = 2.1×10^−186^, β =-0.26) (Table 4, Figure 4). Among 22 proteins with nominal association with MR analysis, an upstream variant (rs4968200) from the FEV1 GWAS colocalized with TNFSF12 protein levels in AGES-Reykjavik (P = 5.6×10^−133^, β = 0.63, RCP = 0.9995).

**Table 4.** Statistics for the variants with strong colocalization (RCP > 0.9)

| gene | SNP | RCP<br>(FEV1 x AGES) | GWAS<br>P-value [beta] | AGES<br>P-value [beta] | deCODE<br>P-value [beta] |
| --- | --- | --- | --- | --- | --- |
| THBS2 | rs3253 <sup>1</sup> | 0.9976 | 1.3x10<sup>-10</sup> [0.02] | 1.4x10<sup>-19</sup> [-0.18] | 2.1x10<sup>-186</sup> [-0.26] |
| THBS2 | rs7756742 <sup>1</sup> | 0.9775 | 6.3x10<sup>-10</sup> [0.02] | 2.8x10<sup>-19</sup> [-0.19] | -- <sup>3</sup> |
| THBS2 | rs7771838 <sup>1</sup> | 0.971 | 7.0x10<sup>-10</sup> [0.02] | 1.8x10<sup>-19</sup> [-0.19] | -- <sup>3</sup> |
| TNFSF12 | rs4968200 <sup>2</sup> | 0.9995 | 4.5x10<sup>-11</sup> [0.02] | 5.6x10<sup>-113</sup> [0.63] | -- <sup>3</sup> |

RCP: Regional Colocalization Probability
<sup>1</sup> Lead variant in FEV1 GWAS is rs3253
<sup>2</sup> Lead variant in FEV1 GWAS is rs4968200
<sup>3</sup> Variant data was not available in DECODE

Finally, all 473 SOMAmers observationally associated with FEV_1_ were included in a MR analysis in the reverse direction, i.e., to evaluate if the changes in SOMAmer levels are downstream of changes in lung function. FEV_1_ was not causally associated with levels of any SOMAmers after adjustment for multiple testing. Data for the 18 SOMAmers that FEV_1_ was nominally associated with (p < 0.05) in the reverse-MR analysis are shown in Table S7.

## Discussion

We present findings from a proteomic analysis of pulmonary function with more candidate protein analytes than previously published to our knowledge (11), highlighting several proteins as strong markers of FEV_1_. Stratification by smoking shows that most associations are driven by ever-smokers. Mendelian randomisation was systematically applied to the candidate markers, revealing proteins whose levels may have a causal effect on lung function. Reverse causation analyses failed to demonstrate that protein level changes associated with lung function occur downstream of the phenotype change, although this could partly be due to insufficient power in the FEV1 GWAS. Among the proteins identified through MR analysis, probabilistic colocalization pointed towards *THBS2* and *TNFSF12* as putative upstream targets for COPD.

Eight proteins were suggested to be causally implicated in lung function based on the MR analysis. However, only three (THBS2, ERO1B and APOM) had consistent direction of effect for the observational and causal estimates. Such discrepancy has been observed when comparing causal and observational estimates (36, 37) for serum proteins. Based on probabilistic quantification, a 3’ UTR variant within *THBS2* was identified as a putative causal variant for FEV1 changes and this lead variant colocalized with THBS2 protein expression in AGES-Reykjavik (Table 4, Figure 4). The effect of this variant was replicated in an independent cohort for THBS2 protein levels (Table 4, Figure 4). Because directions of effects were consistent across the datasets and THBS2 had non-revertible causal association to FEV1, supported by colocalization, THBS2 may be a candidate target for COPD. THBS2 is an extracellular matrix protein that has been implicated in various cardiovascular disorders and is also a candidate biomarker for non-small cell lung cancer (38, 39). *THBS2* is involved in tissue repair and interacts with many different ligands in the extracellular matrix, among them matrix metalloproteases and elastase (40). Although mechanism of *THBS2* needs further experimental validation, our work suggests that protein levels of *THBS2* may be a target for extracellular matrix and regenerative properties associated with COPD. Meanwhile, ERO1B is a disulfide oxidase in the endoplasmic reticulum that is shown to predict survival in pancreatic and pulmonary cancers (41–43) and APOM, an apolipoprotein that is mainly a component of high density lipoproteins and has been associated with COPD severity (44). Genetic variants flanking this gene have been associated with obstructive spirometry measurements (45). However, it must be kept in mind that the association of APOM is based on a single SNP, rs2736163, which is intronic to *PRRC2A*.

Despite not reaching the study threshold for statistical significance, some proteins that were nominally associated in MR analyses are of interest. First, alpha-1-antitrypsin (SERPINA1) is the best-known protein known to cause COPD as severe deficiency of alpha-1-antitrypsin results in obstructive lung disease (46). However, in our data, serum levels of alpha-1-antitrypsin are inversely associated with FEV_1_ in both observational and MR analyses, contrary to what would possibly be expected, although this directionality is known from previous observational analyses of FEV1 and explained by alpha-1-antitrypsin’s role as an acute phase reactant (47). Also, polymorphisms that cause mild or intermediate alpha-1-antitrypsin deficiency are not consistently associated with decreased lung function, suggesting that levels of the protein may only affect lung function below a threshold level (48). Second, TNFSF12 was a protein with a colocalizing pQTL variant with the FEV1 GWAS (Table 4, Figure 4). TNFSF12 is a member of the Tumor Necrosis Factor (TNF) superfamily, of which one key cytokine, TNF-alpha, is a well-known protein target disrupted in COPD patients (49). Despite a concrete association between TNF-alpha levels and COPD in both human and murine models (49–51), TNF-alpha monoclonal antibody was a failure as a COPD therapy (52). This is likely due to COPD being a highly complex inflammatory disease in which single cytokine blockage may not be sufficient for treating the disease. These additional TNF family proteins with MR based causal evidence and probabilistic colocalization may be novel targets for COPD patients impacted by TNF-alpha associated pathobiology. Notable other nominally significant or directionally inconsistent proteins in the findings are matrix metalloproteinase 8 (MMP-8), one of the proteases observationally associated with lung function and implicated in the pathogenesis of COPD (53, 54), TIMP4, an inhibitor of matrix metalloproteinases that has been shown to be upregulated in COPD patients (55) and CD14, levels of which have been shown to be elevated in lungs of smokers (56). While only 8 proteins were significantly associated with FEV_1_ in MR analyses, FEV_1_ was not associated with any proteins in analyses in the reverse direction. The data were therefore unable to support prior causal analyses involving inflammatory markers that have suggested reverse causality of FEV1 with inflammatory markers (18) (Table S6).

The study reveals novel markers with strong observational relations to lung function such as RSPO4, a signalling molecule that is part of the Wnt signalling pathway (57), the tumor marker ALPPL2 (58), the adipokine chemerin (RARRES2), and SVEP1, a protein that is thought to play a role in inflammation in atherosclerosis (59). In addition, the findings validate the observational associations of some of the previously suggested protein markers of FEV_1_, such as SFTPD, fibrinogen, eotaxin and CRP (Table S2) (13), while the associations of other previously suggested biomarkers of FEV_1_ such as AGER were not corroborated in this study (13, 60). Finally, the findings show that proteins that take part in immune responses and extracellular matrix modulation are over-represented among the proteins related to FEV_1_.

This work is subject to a number of limitations. First, the study is based on SOMAmer technology, a relatively novel aptamer-based technology for protein measurements. While many of these SOMAmers have been validated with encouraging results, it has been pointed out that a minority of SOMAmers could be subject to cross-reactivity with related or homologous proteins (20). Second, both the AGES-Reykjavik cohort and the UKBiobank and SpiroMeta Consortium are of European ancestry (27). Therefore, the observed associations could not be generalizable to other populations. Additionally, the AGES-Reykjavik cohort is older than the UKBiobank and SpiroMeta Consortium which could distort comparisons between observational and genetic findings. Third, the measure of lung function used in this paper, FEV_1_, is disproportionally impaired in obstructive lung disease and is a key diagnostic parameter for such diseases. While COPD is the archetypal obstructive lung disease, especially among the elderly, the presence of other respiratory diseases such as asthma could influence the findings, especially in younger cohorts. Fourth, subtle differences in genetic structure between AGES, UKBiobank, and SpiroMeta cohorts is present and may be contributing to the lack of colocalization for the other 6 MR identified proteins. For instance, very few MR instruments used overlapped with 95% credible-sets identified (Supplementary Table 16). In comparisons to UKBioBank, missing variants in AGES-Reykjavik cohort may contribute to colocalization false negatives. Lastly, many causal estimates are directionally inconsistent with observational estimates. While this phenomenon is known from previous proteogenomic studies, its reasons are unclear.

In conclusion, this proteogenomic analysis reveals several proteins that are potentially causally related to lung function, most notably THBS2, ERO1B and APOB.

## Data Availability

All data produced in the present study are available upon reasonable request to the authors

---

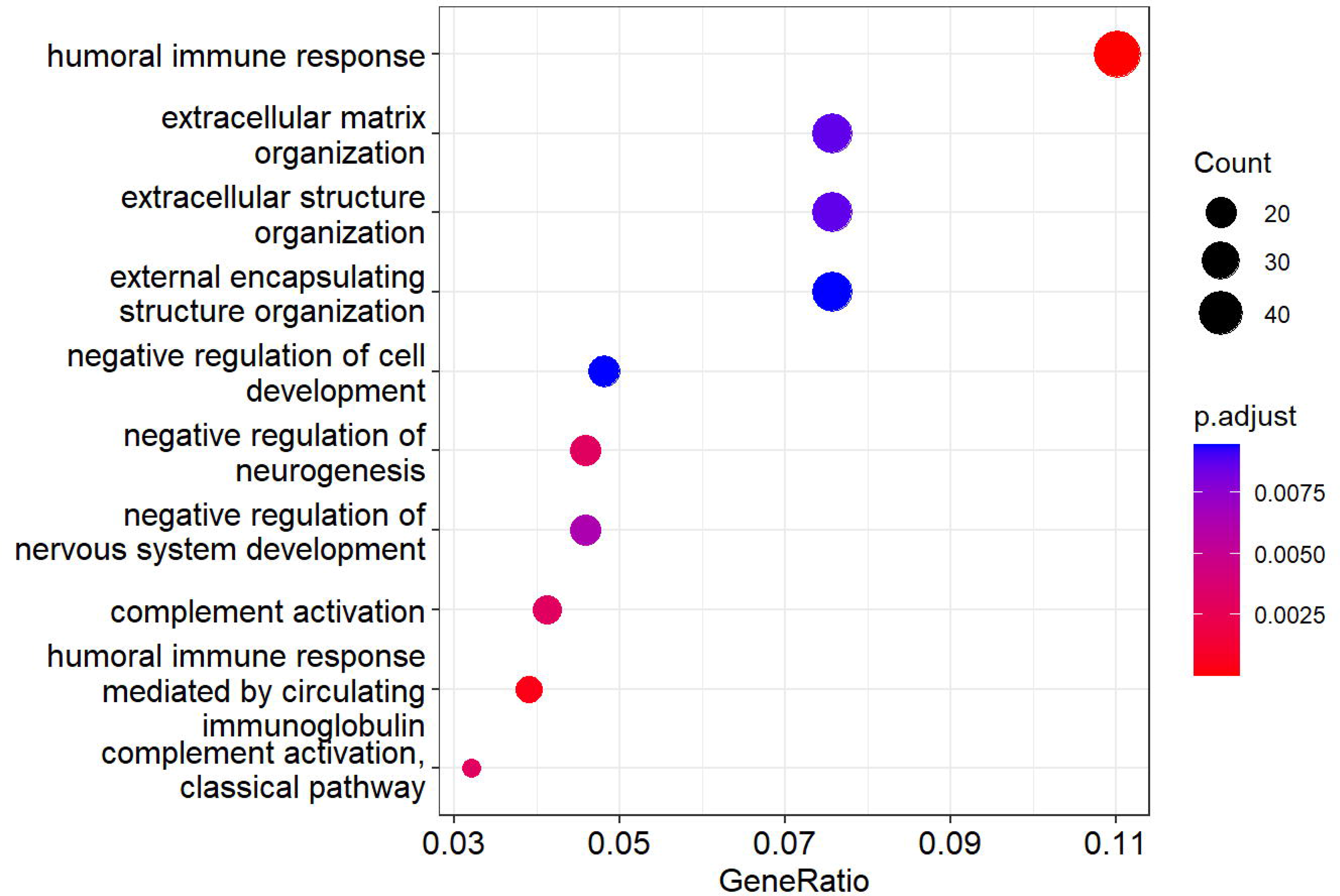

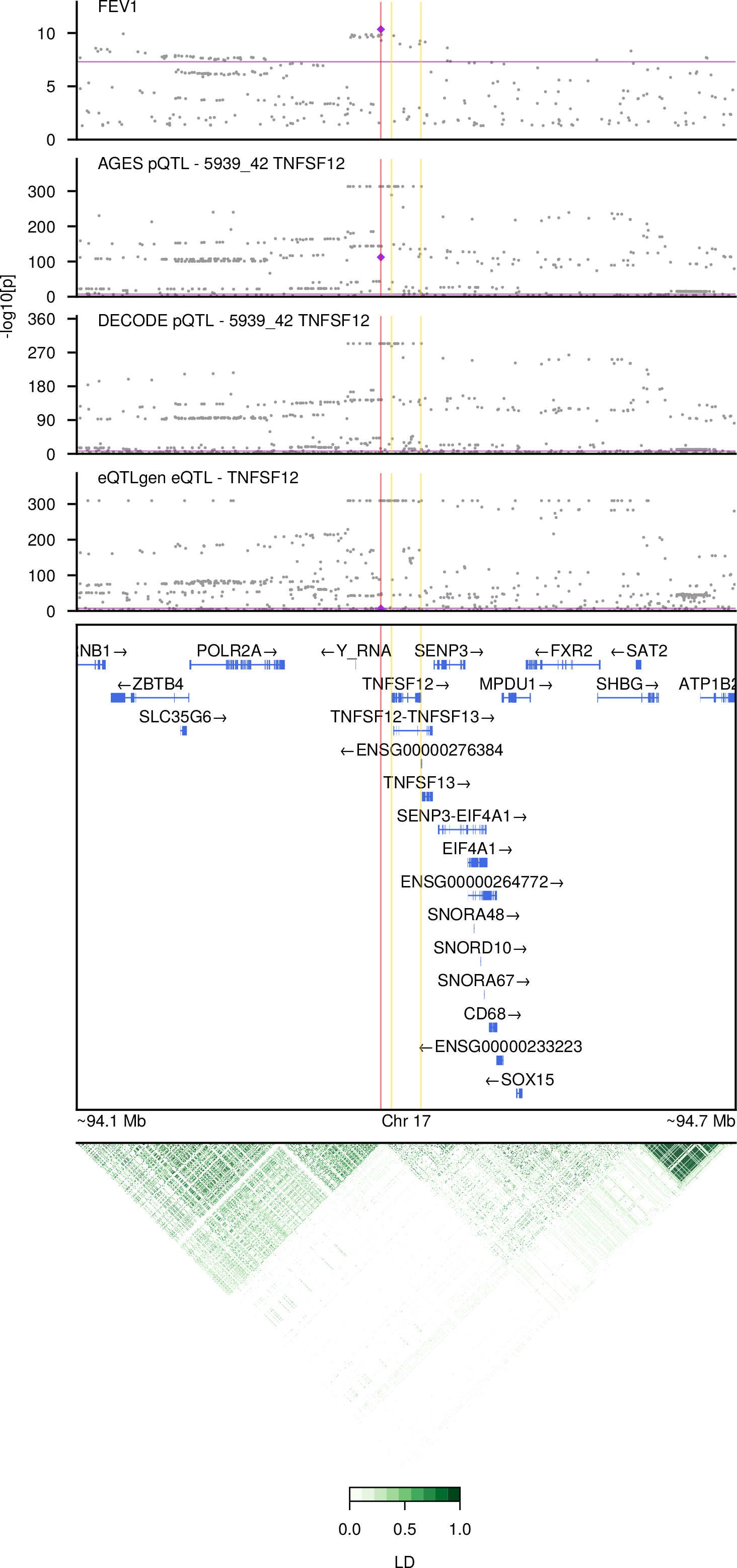

