## Supplement Legends and Tables S2, S3, S6, S7 for "Proteomic associations with forced expiratory volume – a Mendelian randomisation study"

### **Proteomic associations with forced expiratory volume – a Mendelian randomisation study - Supplementary Tables and Figures**

Axelsson GT^1,2^, Jonmundsson T^1^, Woo YJ^3^, Frick EA^1^, Aspelund T^1,5^, Loureiro JJ^3^, Orth AP^4^, Jennings LL^3^ Gudmundsson G^5,6^, Emilsson V^1,5^, Gudmundsdottir V^1,5^, and Gudnason V^1,5^

1: Icelandic Heart Association, Holtasmari 1, IS-201 Kopavogur, Iceland.

2: Landspitali University Hospital, Department of Internal Medicine, 101 Reykjavik, Iceland

3: Novartis Institutes for Biomedical Research, 22 Windsor Street, Cambridge, MA 02139, USA
4: Novartis Institutes for Biomedical Research, 10675 John Jay Hopkins Drive, San Diego, CA 92121, USA

5: Faculty of Medicine, University of Iceland, 101 Reykjavik, Iceland.
6: Landspitali University Hospital, Department of Respiratory Medicine and Sleep, 108 Reykjavik, Iceland

Table S1 - Observational associations of proteins with FEV_1_ (online Excel file)

Table S2 – Observational associations of previously suggested biomarkers of FEV_1_

| **Protein** | **EGS** | **SOMAmer** | **Obs B** | **Obs P** | **Obs FDR** | **Cons** | **Citation** |
| --- | --- | --- | --- | --- | --- | --- | --- |
| GRP78 | HSPA5 | - | - | - | - | - | (60) |
| sCD163 | CD163 | 5028_59 | 0.002 | 0.87 | 0.95 | * | (60) |
| CC16 | SCGB1A1 | 10569_28 | -0.025 | 0.07 | 0.27 | * | (13) |
| SP-D | SFTPD | 4414_69 | -0.05 | 0.0002 | 0.005 | Yes | (13) |
| sRAGE | AGER | 4125_52 | -0.01 | 0.33 | 0.61 | * | (13) |
| CRP | CRP | 4337_49 | -0.06 | 1.2×10^-7^ | 2.6×10^-5^ | Yes | (13) |
| Fibrinogen | FGA | 4907_56 | -0.04 | 0.0008 | 0.015 | Yes | (13) |
| Fibrinogen | FGA | 2796_62 | -0.005 | 0.67 | 0.85 | * | (13) |
| IL-6 | IL6 | 4673_13 | -0.04 | 5.3×10^-4^ | 0.012 | Yes | (61) (62) |
| IL-6 | IL6 | 2573_20 | -0.05 | 9.6×10^-5^ | 0.003 | Yes | (61) (62) |
| P-selectin | SELP | 4154_57 | -0.014 | 0.25 | 0.53 | * | (61) |
| Eotaxin | CCL11 | 5301_7 | -0.05 | 0.0002 | 0.005 | Yes | (62) |
| IFN-γ | IFNG | 2989_17 | -0.008 | 0.58 | 0.80 | * | (62) |
| IFN-γ | IFNG | 14147_50 | -0.008 | 0.55 | 0.78 | * | (62) |
| IL-10 | IL10 | 2773_50 | -0.0015 | 0.92 | 0.97 | * | (62) |
| IL-10 | IL10 | 13723_6 | 0.02 | 0.09 | 0.29 | * | (62) |
| IL-2 | IL2 | 3070_1 | 0.015 | 0.25 | 0.52 | * | (62) |
| IL-8 | CXCL8 | 3447_64 | 0.003 | 0.84 | 0.94 | * | (62) |
| TNF-α | TNF | 5936_53 | -0.010 | 0.45 | 0.71 | * | (62) |
| TNF-α | TNF | 5692_79 | -0.021 | 0.10 | 0.31 | * | (62) |

Adjusted for sex, age, age squared, height and height squared.

-: No data available
*: Not applicable as observational analysis in AGES-Reykjavik were not significant

Table S3 – Observational associations of proteins with FEV_1_ stratified by ever-smoking

|  |  | **EVER-SMOKERS**  **(n = 969)** | | | **NEVER-SMOKERS**  **(n = 662)** | | |
| --- | --- | --- | --- | --- | --- | --- | --- |
| **SOMA** | **EGS** | **β** | **95% CI** | **P** | **β** | **95% CI** | **P** |
| 8464_31_3 | RSPO4 | -0.106 | -0.141 - -0.071 | 4.83×10^-9^ | -0.029 | -0.067 - 0.009 | 0.135 |
| 7813_6_3 | ALPPL2 | -0.092 | -0.125 - -0.059 | 5.73×10^-8^ | -0.009 | -0.047 - 0.029 | 0.642 |
| 3079_62_2 | RARRES2 | -0.11 | -0.147 - -0.072 | 1.19×10^-8^ | -0.034 | -0.073 - 0.004 | 0.083 |
| 11178_21_3 | SVEP1 | -0.085 | -0.121 - -0.049 | 4.98×10^-6^ | -0.081 | -0.116 - -0.047 | 4.52×10^-6^ |
| 3216_2_2 | PIGR | -0.077 | -0.11 - -0.044 | 6.76×10^-6^ | -0.034 | -0.069 - 0.002 | 0.062 |
| 11109_56_3 | SVEP1 | -0.083 | -0.119 - -0.047 | 7.09×10^-6^ | -0.082 | -0.117 - -0.047 | 5.06×10^-6^ |
| 2292_17_4 | C9 | -0.095 | -0.129 - -0.061 | 5.65×10^-8^ | -0.035 | -0.068 - -0.001 | 0.042 |
| 12549_33_3 | HPGDS | 0.088 | 0.054 - 0.123 | 6.07×10^-7^ | 0.043 | 0.011 - 0.076 | 9.73×10^-3^ |
| 6379_62_3 | ADAMTSL2 | -0.076 | -0.112 - -0.039 | 5.62×10^-5^ | -0.064 | -0.101 - -0.027 | 6.74×10^-4^ |
| 9191_8_3 | TFF2 | -0.095 | -0.129 - -0.061 | 3.84×10^-8^ | -0.007 | -0.043 - 0.028 | 0.681 |
| 8841_65_3 | CILP2 | 0.087 | 0.053 - 0.121 | 6.12×10^-7^ | 0.029 | -0.004 - 0.062 | 0.087 |
| 8323_163_3 | TFF3 | -0.097 | -0.134 - -0.059 | 4.36×10^-7^ | -0.031 | -0.066 - 0.003 | 0.073 |
| 6390_18_3 | NPS | 0.088 | 0.051 - 0.124 | 2.48×10^-6^ | 0.052 | 0.017 - 0.086 | 3.18×10^-3^ |
| 2211_9_6 | TIMP1 | -0.122 | -0.167 - -0.077 | 1.18×10^-7^ | -0.024 | -0.07 - 0.022 | 0.3 |
| 13722_105_3 | C9 | -0.083 | -0.117 - -0.05 | 1.37×10^-6^ | -0.033 | -0.067 - 0 | 0.049 |
| 6605_17_3 | IGFALS | 0.078 | 0.044 - 0.113 | 9.03×10^-6^ | 0.034 | 0.002 - 0.066 | 0.041 |
| 12707_26_3 | DPYSL3 | 0.079 | 0.044 - 0.114 | 8.26×10^-6^ | 0.021 | -0.013 - 0.054 | 0.222 |
| 5728_60_3 | FCRL1 | 0.087 | 0.054 - 0.12 | 3.72×10^-7^ | 0.016 | -0.018 - 0.049 | 0.368 |
| 6075_61_3 | HEXB | 0.078 | 0.041 - 0.115 | 3.47×10^-5^ | 0.048 | 0.015 - 0.082 | 5.17×10^-3^ |
| 4496_60_2 | MMP12 | -0.068 | -0.102 - -0.034 | 1.06×10^-4^ | -0.033 | -0.068 - 0.002 | 0.065 |
| 6225_3_3 | PRSS8 | 0.081 | 0.044 - 0.119 | 2.25×10^-5^ | 0.054 | 0.02 - 0.088 | 1.99×10^-3^ |
| 4337_49_2 | CRP | -0.061 | -0.093 - -0.029 | 1.93×10^-4^ | -0.05 | -0.082 - -0.018 | 2.31×10^-3^ |
| 10620_21_3 | MSMB | -0.091 | -0.127 - -0.054 | 1.08×10^-6^ | -0.014 | -0.05 - 0.022 | 0.443 |
| 2609_59_2 | CST3 | -0.095 | -0.133 - -0.058 | 8.23×10^-7^ | -0.034 | -0.073 - 0.005 | 0.084 |
| 8885_6_3 | CACNA2D3 | 0.087 | 0.052 - 0.122 | 1.55×10^-6^ | 0.003 | -0.031 - 0.037 | 0.863 |

Results are shown for the 25 proteins with the most significant associations with FEV1 in the whole cohort.

Table S4 – Results of over-representation analyses of GO terms related to genes annotated to FEV1 associated SOMAmers (online Excel file)

Table S5 – Genetic instruments used in MR analyses for SOMAmers significantly associated with FEV_1_ (online Excel file)

Table S6 – MR associations of proteins previously suggested to be causally related to FEV_1_

| **Protein** | **EGS** | **SOMAmer** | **MR B** | **MR P** | **MR FDR** | **Rev MR B** | **Rev MR P** | **Rev MR FDR** | **Citation** |
| --- | --- | --- | --- | --- | --- | --- | --- | --- | --- |
| SP-D | SFTPD | 4414_69 | - | - | - | 0.12 | 0.07 | 0.95 | (13) |
| CRP | CRP | 4337_49 | 0.0002 | 0.99 | 1.0 | -0.09 | 0.13 | 0.95 | (13) |
| Fibrinogen | FGA | 4907_56 | -0.003 | 0.86 | 0.98 | -0.09 | 0.16 | 0.95 | (13) |
| IL-6 | IL6 | 4673_13 | - | - | - | -0.0003 | 1 | 1 | (61) (62) |
| IL-6 | IL6 | 2573_20 | - | - | - | -0.03 | 0.67 | 0.95 | (61) (62) |
| Eotaxin | CCL11 | 5301_7 | -0.04 | 0.11 | 0.49 | -0.02 | 0.77 | 0.97 | (62) |

-: No data available

Table S7 – Results of Mendelian randomisation analysis of the association of FEV_1_ with proteins

| **SOMA** | **Protein** | **nSNP** | **Β - MR** | **SE** | **P - MR** | **FDR P - MR** | **Β - Obs** | **P - Obs** | **FDR P - Obs** |
| --- | --- | --- | --- | --- | --- | --- | --- | --- | --- |
| *9282_12_3* | *CRISP2* | *366* | *0.195* | *0.056* | *4.94e-04* | *0.234* | *0.074* | *3.36×10^-7^* | *5.55×10^-5^* |
| *5034_79_1* | *PRSS2* | *366* | *-0.155* | *0.061* | *0.012* | *0.948* | *-0.042* | *6.43×10^-4^* | *0.013* |
| 12395_86_3 | DARS2 | 366 | 0.144 | 0.06 | 0.015 | 0.948 | -0.037 | 3.09×10^-3^ | 0.036 |
| *14260_112_3* | *NET1* | *366* | *-0.142* | *0.059* | *0.017* | *0.948* | *-0.039* | *2.29×10^-3^* | *0.03* |
| *7994_41_3* | *ERO1B* | *366* | *-0.142* | *0.061* | *0.02* | *0.948* | *-0.036* | *3.78×10^-3^* | *0.042* |
| *3324_51_1* | *LY9* | *366* | *0.139* | *0.06* | *0.02* | *0.948* | *0.042* | *1.08×10^-3^* | *0.017* |
| 12740_55_3 | FEV | 366 | 0.139 | 0.06 | 0.021 | 0.948 | -0.042 | 6.37×10^-4^ | 0.013 |
| *3505_6_2* | *LTA* | *365* | *0.137* | *0.06* | *0.023* | *0.948* | *0.052* | *6.40×10^-5^* | *2.68×10^-3^* |
| *6380_23_3* | *MRPL58* | *365* | *-0.146* | *0.064* | *0.024* | *0.948* | *-0.057* | *8.24×10^-6^* | *5.80×10^-4^* |
| *2575_5_5* | *LEP* | *366* | *-0.119* | *0.053* | *0.025* | *0.948* | *-0.044* | *2.76×10^-3^* | *0.034* |
| *4891_50_1* | *GCG* | *366* | *-0.135* | *0.061* | *0.026* | *0.948* | *-0.038* | *2.79×10^-3^* | *0.034* |
| *8476_11_3* | *CHGA* | *364* | *-0.131* | *0.059* | *0.026* | *0.948* | *-0.055* | *1.50×10^-5^* | *9.18×10^-4^* |
| *2867_52_2* | *AKT1* | *365* | *0.132* | *0.06* | *0.028* | *0.948* | *0.055* | *1.60×10^-5^* | *9.56×10^-4^* |
| *6236_51_3* | *CTHRC1* | *366* | *-0.105* | *0.051* | *0.037* | *0.948* | *-0.047* | *9.85×10^-4^* | *0.017* |
| 10754_113_3 | PROK2 | 364 | 0.135 | 0.065 | 0.038 | 0.948 | -0.038 | 2.32×10^-3^ | 0.03 |
| *2950_57_2* | *IGFBP4* | *366* | *-0.122* | *0.06* | *0.041* | *0.948* | *-0.042* | *1.22×10^-3^* | *0.019* |
| *11178_21_3* | *SVEP1* | *366* | *-0.117* | *0.058* | *0.042* | *0.948* | *-0.085* | *1.67×10^-10^* | *1.72×10^-7^* |
| *13098_93_3* | *VEGFD* | *366* | *-0.112* | *0.055* | *0.044* | *0.948* | *-0.048* | *2.66×10^-4^* | *7.31×10^-3^* |

Shown are data for observationally significant SOMAmers that had nominally significant associations using Mendelian randomisation. Observational data are adjusted for sex, age, age squared, height and height squared.

*Italic* = Consistent in direction between MR analyses and observational analyses

#### Supplementary sensitivity analyses (online Excel files)

Table S8 – Observational associations with FEV_1_, corrected for smoking

Table S9 – Observational associations with FEV_1_, corrected for smoking and eGFR

Table S10 – Observational associations with FVC, corrected for smoking

Table S11 – Observational associations with FVC, corrected for smoking and eGFR

Table S12 – Observational associations with FEV_1_/FVC, corrected for smoking

Table S13 – Observational associations with FEV_1_/FVC, corrected for smoking and eGFR

Table S14 – Observational associations with COPD, corrected for smoking

Table S15 – Observational associations with COPD, corrected for smoking and eGFR

Table S16 – Overlap between colocalization credible sets and MR instruments

Figure S1 – Results of over-representation analyses of GO terms associated with genes annotated to FEV1 associated SOMAmers

Figure S2. Colocalization plot for TNFSF12
